# Prevalence of laboratory-related musculoskeletal disorders among biomedical scientists

**DOI:** 10.1101/2021.06.04.21258372

**Authors:** Nasar Alwahaibi, Mallak Al Sadairi, Ibrahim Al Abri, Samira Al Rawahi

## Abstract

**Background:** Laboratory–related musculoskeletal disorders (LMSDs) are injuries resulted from working in the laboratory. Biomedical scientists (BMSs) play an important role in any health care system. However, they are at high risk of exposure to LMSDs. This study aimed to estimate the prevalence and the associated risk factors of LMSDs among this group of healthcare professionals.

**Methods:** A cross-sectional survey (Nordic musculoskeletal) was used to estimate the prevalence of LMSDs among BMSs. Data were analyzed using Statistical Package for Social Science software version 25. Chi-square was performed to find the significant association between LMSDs and different risk factors.

**Results:** The study included 83 BMSs. Females represented 63.9% and 36.1% were in the age group of 35–44. The overall prevalence of LMSDs was 77.1%. The most prevalent LMSDs were neck, shoulders, and lower back with 50.6%, 49.4%, and 43.4%, respectively. Neck complaints and upper back complaints were found statistically significant with the female gender. Shoulders complaints were associated with pipetting and microscopy. Lower back complaints were associated with pipetting and heavy work at home. A total of 65.57% of BMSs had irregular symptoms of LMSDs, 54.10% experienced moderate pain due to these symptoms, and 44.26% had symptoms that persisted from hours to days.

**Conclusion:** The study found that the prevalence of LMSDs among BMSs was high. Good knowledge, attitude, practice, and training of ergonomics may minimize the prevalence of LMSDs among BMSs.

## Introduction

Musculoskeletal disorders (MSDs), known as cumulative trauma disorders or repetitive stress injuries, are injuries to muscles, nerves, tendons, ligaments, joints, cartilage, and spinal discs (1). Whereas work-related musculoskeletal disorders (WMSDs) are conditions in which the work environment and performance of work contribute significantly to the condition, and/or the condition is made worse or persists longer due to work conditions (2). To the best of our knowledge, we are the first to introduce the term laboratory-related musculoskeletal disorders (LMSDs) and can be defined simply as injuries resulted from working in the laboratory.

Biomedical scientists (BMSs) play an important role in any health care institutions. They work in medical laboratories, where specimens are prepared and processed for the detection, diagnosis and treatment of various diseases. BMSs are professionals that identify pathogenic microorganisms in microbiology, measure chemicals in biochemistry, match blood samples in hematology, help to diagnose cancer samples in histopathology, recognize antigen molecules in immunology, detect gene variants in genetics, and others. In medical laboratory, they are exposed to repetitive activities such as microscopy, microtomy, pipetting, and mostly work on standing positions. With the time, repetitive motion can affect muscles, joints, tendons and nerves (3). The long-term and severe LMSDs can highly affect the productivity of the work and shortness the work duration, therefore this will eventually lead to occupational disability and cause a major health challenge for individuals and healthcare systems around the world (4). About 33% of all sickness absence of health care professionals result from MSDs (5). MSDs are one of the common health problems among all health professionals (6). In addition, about 30% of people worldwide have at least MSDs (7).

Few studies are available for the ergonomic hazard which can lead to LMSDs (8). Ergonomic is simply the balance between the work requirement and the capacity of the worker (9). In addition, MSDs studies on certain professionals such as dentists, nurses and surgeons are well covered, however, data on the prevalence of LMSDs among BMSs is scarce. Hence, this study aimed to evaluate the prevalence of LMSDs among BMSs and assess the risk factors associated with BMSs.

## Methods

This is a cross-sectional observational study that was conducted in the year of 2020 at the Sultan Qaboos University Hospital and Sultan Qaboos University. The study is ethically approved from the Medical Research Ethics Committee, College of Medicine and Health Sciences, SQU (SQU-EC/095/2020). Preceding the study, a detailed procedure of the study was explained, a voluntary consent form was also signed by each participant before the study. BMSs with occupational or non-occupational accidents that affected their musculoskeletal system were excluded from the study.

The sample size was calculated using the infinite population sample size formula: n = {N × Z^2^ × p × (1–p)} / {d^2^ × (N–1) + Z^2^× p × (1 – p) where N : Population size (The total number of BMS working at SQU and SQUH) = 170, Z (Standard value with confidence level 95%) =1.96, d(Permissible error on either side) = 10%, p (Proportion of the characteristic under the study). At the present, there is no available information on the proportion of MSDs among BMS. Therefore, the *P*-value was obtained from a pilot study which was conducted among 20 participants who were fulfilled the research criteria. Those who participated in the pilot study were excluded from the study. The *P*-value was found equal to 63.5%. The sample size was calculated as 97. After sampling, the number was increased 15% to avoid non-response or inappropriately filled questions and that number was randomly selected from the overall population.

The survey included two main sections. The first section was used to obtain data about socio-demographic characteristics such as age, gender, weight, height, nationality, specialty, educational level, nature of the job, and the number of experience years. The second section was from a previously validated Nordic musculoskeletal questionnaire (10). This section was used to assess the prevalence of MSDs among the participants. The first two questions were about the health status of the participants and whether they had any occupational or non-occupational accidents that affected their musculoskeletal system. Therefore, those who answered yes in these two questions were excluded because they did not fulfill the research criteria. The participants were asked in this section if they experienced any pain at different nine anatomical regions of the body, specifically at: neck, shoulder, upper back, elbows, lower back, wrists/hands, hips/thighs, knees, and feet/ankles in the last year preceding the survey and also in the last seven days before the study. Further, the participants were asked in this section if they visited a physiotherapist or prevented from doing works due to any pain in the same nine regions mentioned above. There was a diagram to show these body sites for clarification to the participants. The section also includes questions about the intensity, frequency, and duration of the pain related to any musculoskeletal injuries.

The data were analyzed using Statistical Package for Social Science (SPSS) version 25 software. Frequencies and percentages were used to represent the categorical data such as age, gender, and specialty. Continuous data were presented as mean and standard deviation. A Chi-square test was performed to measure the significant association between different risk factors and the prevalence of LMSDs among the BMSs. The *P*-value was considered significant if it was equal to or less than 0.05.

## Results

A total of 114 BMSs participated in the study. However, 31 BMSs were excluded because they did not fulfil the research criteria. Therefore a total of 83 BMSs were included in this study. The vast majority were females (63.9%) and 75.9% were married. The majority of the BMSs (36.1%) were in the age group of 35-44. Their average body mass index (BMI) was 25.3 (±4.6). More than half of the participants (53.0%) did regular physical exercises. Almost all the participants are right-handed (94.0%). More than half (73.2%) of the participants used to work while they were standing. A total of 60.2% BMSs were sometimes stressed at work, 50.0% were sometimes working overtime, and 79.0% did not have heavy work at home (Table 1).

**Table1:**
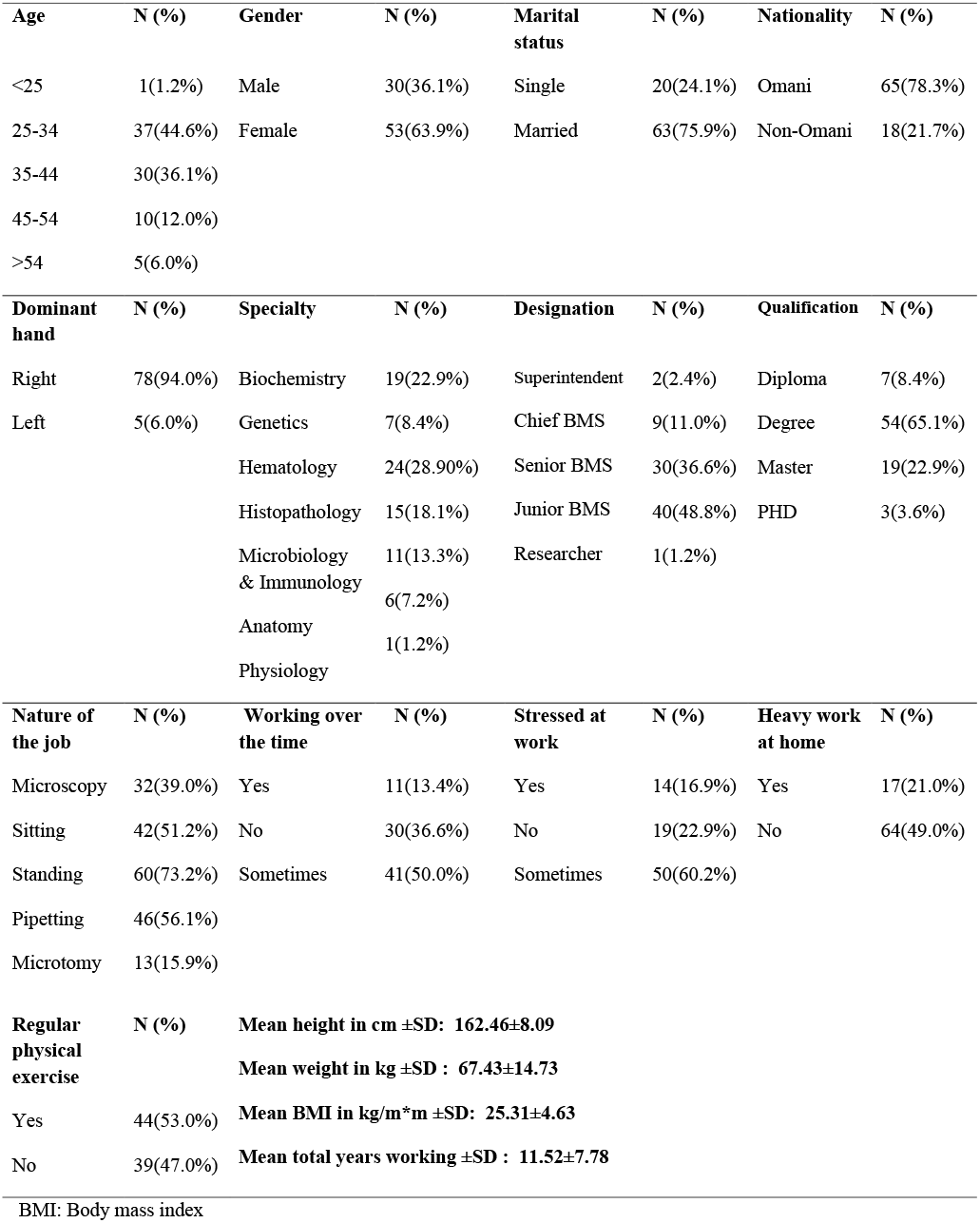
Demographic data of biomedical scientists

The overall prevalence of LMSDs among BMSs in the last 12 months in any parts of the body was 77.1%. Neck complaint was the highest; 50.6% and 25.3% in the last 12 months and 7 days respectively. Followed by shoulders complaint 49.4% and 16.9%, lower back complaint 43.4% and 15.7%, upper back compliant 34.9% and 14.5%, knees compliant 30.1% and 8.4%, ankle or feet 26.5% and 4.8%, wrists or hands 24.1 % and 4.8% respectively. Elbows complaint was the lowest in both periods, 9.6% and 2.4% followed by thighs or hip complaints, 10.8%, and 3.6% respectively (Figure 1). A total of 65.57% of BMSs had irregular symptoms of LMSDs, 54.10% experienced moderate pain due to these symptoms, and 44.26% had symptoms that persisted from hours to days (Figure 2).

**Figure 1:**
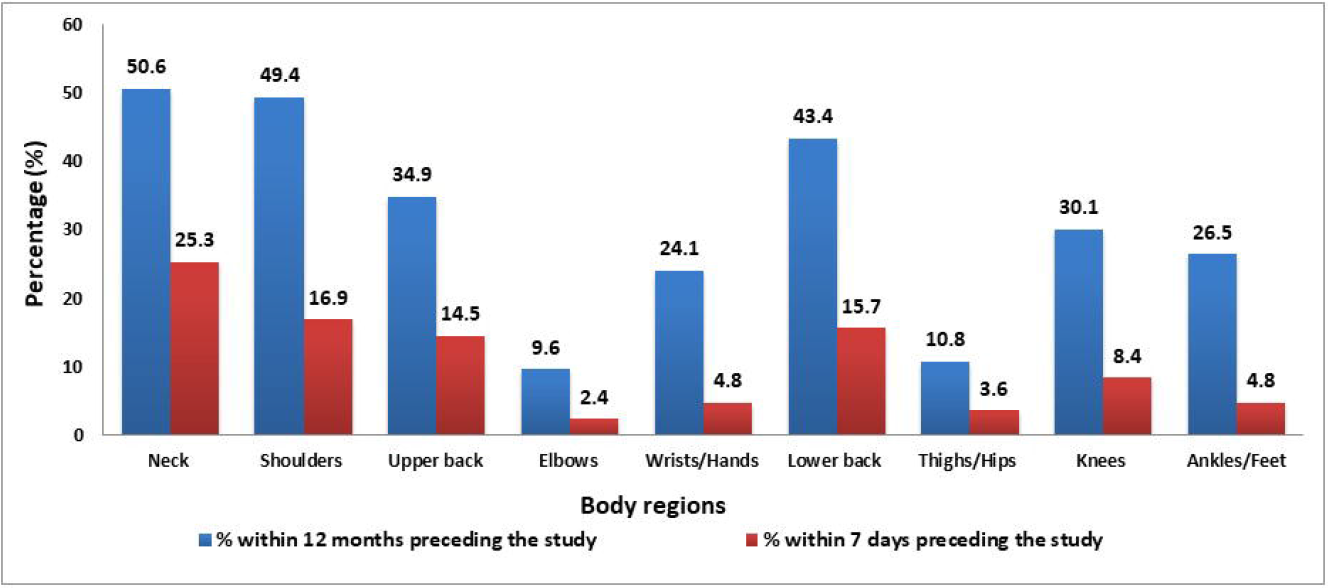
Comparison between the prevalence of LMSDs among BMSs in the last 7 days and 12 months preceding the study.

**Figure 2:**
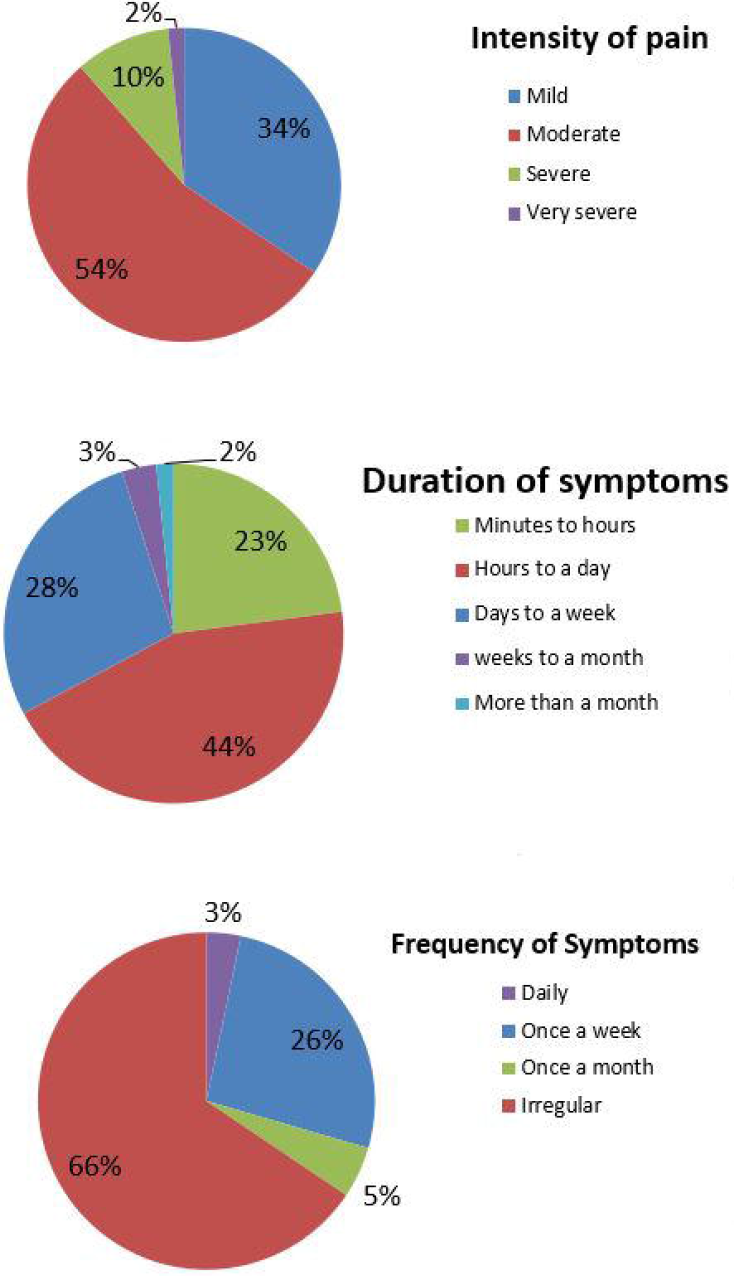
Pain and symptoms characteristics of the LMSDs experienced by BMSs working within 12 months preceding the study.

There was no significant association between age and marital status and LMSDs among BMSs. However, there was a significant association between the neck and upper back complaints with the female gender (Table 2). In addition, no significant association was found between nationality, designation and degree of qualification with the LMSDs among the participants. None of the complaints were significantly associated with dominant hand, BMI, and regular physical exercises. (Table 3). Heavy work at home was significantly associated with a lower back complaint. There was no significant association between working overtime and stressed at work with LMSDs in this study (Table 4). Regarding the nature of the job, shoulders complaint was significantly associated with microscopy and pipetting. Lower back complaints show a significant association with pipetting (Table 5).

**Table 2:** Prevalence and association of LMSDs with age, gender, and nationality of BMSs within 12 months.

| Body regions | Risk Factors |  |  |  |  |  |  |  |  |  |  |  |
| --- | --- | --- | --- | --- | --- | --- | --- | --- | --- | --- | --- | --- |
|  | Age N (%) |  |  |  |  | Gender N (%) |  |  |  | Marital status N (%) |  |  |
|  | 25 | 25-34 | 35-44 | 44-54 | >54 | P-value | Female | Male | P-value | Married | Single | P-value |
| Neck | 1(100%) | 22(59.5%) | 16(53.3%) | 3(30.0%) | 0(0.0%) | 0.060 | 33(62.3%) | 9 (30.0%) | **0.009** | 28(44.4%) | 14(70.0%) | 0.083 |
| Shoulders | 0(0.0%) | 20(54.1%) | 16(53.3%) | 4(40.0%) | 1(20.0%) | 0.468 | 30(56.6%) | 11(36.7%) | 0.129 | 29(46.0%) | 12(60.0%) | 0.405 |
| Upper back | 0(0.0%) | 18(48.6%) | 8(26.7%) | 3(30.0%) | 0.0(0.0%) | 0.121 | 24(45.3%) | 5(16.7%) | **0.017** | 23(36.5%) | 6(30.0%) | 0.793 |
| Elbows | 0(0.0%) | 4(10.8%) | 2(6.7%) | 2(20.0%) | 0(0.0%) | 0.693 | 6(11.3%) | 2(6.7%) | 0.762 | 6(9.5%) | 2(10.0%) | 1.000 |
| Wrists/hands | 1(100%) | 11(29.7%) | 6(20.0%) | 2(20.0%) | 0(0.0%) | 0.219 | 13(24.5%) | 7(23.3%) | 1.000 | 12(19.0%) | 8(40.0%) | 0.108 |
| Lower back | 0(0.0%) | 19(51.4%) | 13(43.3%) | 1(10.0%) | 3(60.0%) | 0.146 | 26(49.1%) | 10(33.3%) | 0.247 | 26(41.3%) | 10(50.0%) | 0.669 |
| Hips/ thighs | 0(0.0%) | 3(8.1%) | 4(13.3%) | 2(20.0%) | 0(0.0%) | 0.722 | 6(11.3%) | 3(10.0%) | 1.000 | 7(11.1%) | 2(10.0%) | 1.000 |
| Knees | 0(0.0%) | 14(37.8%) | 6(20.0%) | 3(30.0%) | 2(40.0%) | 0.530 | 8(26.7%) | 17(32.1%) | 0.789 | 17(27.0%) | 8(40.0%) | 0.409 |
| Ankles/feet | 1(100%) | 11(29.7%) | 7(23.3%) | 3(30.0%) | 0(0.0%) | 0.288 | 7(23.3%) | 15(28.3%) | 0.815 | 13(20.6%) | 9(45.0%) | 0.063 |

**Table 3:** Prevalence and association of LMSDs with dominant hand, body mass index, and physical exercises of BMSs within 12 months.

| Body regions | Risk factors |  |  |  |  |  |  |  |  |  |  |
| --- | --- | --- | --- | --- | --- | --- | --- | --- | --- | --- | --- |
|  | Dominant hand N (%) |  |  | BMI N (%) |  |  |  |  | Physical exercises N (%) |  |  |
|  | Right | Left | *P*-value | Underweigh<br>t | Normal | Overweight | Obese | *P*-<br>value | Yes | No | *P*-value |
| Neck | 41(52.6%) | 1(20.0%) | 0.342 | 3(100.0%) | 16(47.1%) | 17(54.8%) | 4(44.4%) | 0.333 | 21(47.7%) | 21(53.8%) | 0.763 |
| Shoulder | 39(50.0%) | 2(40.0%) | 1.000 | 1(33.3%) | 20(58.8%) | 15(48.4%) | 3(33.3%) | 0.491 | 20(45.5%) | 21(53.8%) | 0.587 |
| Upper back | 29(37.2%) | 0(0.0%) | 0.228 | 3(100.0%) | 12(35.3%) | 8(25.8%) | 4(44.4%) | 0.070 | 13(29.5%) | 16(41.0%) | 0.387 |
| Elbows | 7(9.0%) | 1(20.0%) | 0.977 | 0(0.0%) | 4(11.8%) | 4(12.9%) | 0(0.0%) | 0.643 | 4(9.1%) | 4(10.3%) | 1.000 |
| Wrists<br>&hands | 20(25.65) | 0(0.0%) | 0.447 | 2(66.7%) | 8(23.5%) | 9(29.0%) | 0(0.0%) | 0.105 | 10(22.7%) | 10(25.6%) | 0.958 |
| Lower back | 35(44.9%) | 1(20.0%) | 0.534 | 2(66.7%) | 15(44.1%) | 11(35.5%) | 5(55.6%) | 0.573 | 19(43.2%) | 17(43.6%) | 1.000 |
| Hips/thighs | 9(11.5%) | 0(0.0%) | 0.950 | 0(0.0%) | 2(5.9%) | 6(19.4%) | 0(0.0%) | 0.186 | 5(11.4%) | 4(10.3%) | 1.000 |
| Knees | 25(32.1%) | 0(0.0%) | 0.321 | 1(33.3%) | 10(29.4%) | 8(25.8%) | 5(55.6%) | 0.397 | 12(27.3%) | 13(33.3%) | 0.718 |
| Ankles/feet | 21(26.9%) | 1(20.0%) | 1.000 | 1(33.3%) | 7(20.6%) | 11(35.6%) | 2(22.2%) | 0.574 | 12(27.3%) | 10(25.6%) | 1.000 |

BMI: Body mass index

**Table 4:** Prevalence and association of LMSDs with the working overtime, heavy work at home, and stressed at work among the BMSs within 12 months.

| Body regions | Risk factors |  |  |  |  |  |  |  |  |  |  |
| --- | --- | --- | --- | --- | --- | --- | --- | --- | --- | --- | --- |
|  | Working over the time N (%) |  |  |  | Heavy work at home N (%) |  |  | Stressed at work N (%) |  |  |  |
|  | Yes | No | Sometimes | *P*- value | Yes | No | *P*- value | Yes | No | Sometimes | *P*- value |
| **Neck** | 5(54.5%) | 14(46.7%) | 21(51.2%) | 0.883 | 9(52.9%) | 32(50.0%) | 1.000 | 10(71.4%) | 7(36.8%) | 25(50.0%) | 0.144 |
| **Shoulder** | 5(45.5%) | 16(53.5%) | 19(46.3%) | 0.821 | 9(52.9%) | 31(48.4%) | 0.954 | 8(57.1%) | 6(31.6%) | 27(54.0%) | 0.205 |
| **Upper back** | 5(45.5%) | 10(33.3%) | 13(31.7%) | 0.690 | 8(47.1%) | 20(31.3%) | 0.352 | 4(28.6%) | 4(21.1%) | 21(42.0%) | 0.228 |
| **Elbows** | 1(9.1%) | 2(6.7%) | 4(9.8%) | 0.897 | 3(17.6%) | 4(6.3%) | 0.371 | 2(14.3%) | 1(5.3%) | 5(10.0%) | 0.680 |
| **Wrists & hands** | 2(18.2%) | 8(26.7%) | 9(22.0%) | 0.821 | 4(23.5%) | 14(21.9%) | 1.000 | 2(14.3%) | 4(21.1%) | 14(28.0%) | 0.535 |
| **Lower back** | 6(54.5%) | 11(36.7%) | 18(43.9%) | 0.577 | 12(70.6%) | 23(35.9%) | **0.022** | 8(57.1%) | 5(26.3%) | 23(46.0%) | 0.176 |
| **Hips/thighs** | 1(9.1%) | 4(13.3%) | 3(7.3%) | 0.698 | 3(17.6%) | 5(7.8%) | 0.583 | 1(7.1%) | 0(0.0%) | 8(16.0%) | 0.143 |
| **Knees** | 3(27.3%) | 9(30.0%) | 12(29.3%) | 0.986 | 8(47.1%) | 16(25.0%) | 0.496 | 4(28.6%) | 5(26.3%) | 16(32.0%) | 0.891 |
| **Ankles/feet** | 4(36.4%) | 7(23.3%) | 10(24.4%) | 0.677 | 6(35.3%) | 15(23.4%) | 0.141 | 3(21.4%) | 3(18.5%) | 16(32.0%) | 0.353 |

**Table 5:** Prevalence and association of LMSDs with the nature of the job of BMSs within 12 months.

| Body regions | Nature of the job N (%) |  |  |  |  |  |  |  |  |  |
| --- | --- | --- | --- | --- | --- | --- | --- | --- | --- | --- |
|  | Microscopy | *P- value* | Sitting | *P- value* | Standing | *P-value* | Pipetting | *P- value* | Microtomy | *P-value* |
| Neck | 20(62.5%) | 0.159 | 19(45.2%) | 0.374 | 30(50.0%) | 0.908 | 27(58.7) | 0.191 | 7(53.8%) | 1.000 |
| Shoulder | 21(56.6%) | **0.042** | 21(50.0%) | 1.000 | 29(48.3%) | 0.803 | 28(26.9%) | **0.045** | 5(58.5%) | 0.545 |
| Upper back | 15(46.9%) | 0.132 | 13(31.0%) | 0.532 | 23(28.3%) | 0.504 | 19(41.3%) | 0.299 | 5(83.5%) | 1.000 |
| Elbow | 6(18.8%) | 0.070 | 3(7.1%) | 0.656 | 5(8.3%) | 0.766 | 3(6.5%) | 0.495 | 4(30.8%) | *NA |
| Wrists/Hands | 9(28.1%) | 0.714 | 8(19.0) | 0.730 | 16(26.7%) | 0.615 | 12(28.1%) | 0.884 | 5(38.5%) | 0.349 |
| Lower back | 14(43.8%) | 1.000 | 20(47.6%) | 0.736 | 28(46.7%) | 0.561 | 25(24.3%) | **0.054** | 7(53.8%) | 0.629 |
| Hips/Thighs | 3(9.4%) | 0.993 | 3(7.1%) | 0.433 | 6(10.0%) | 0.647 | 5(10.9%) | 1.000 | 2(15.4%) | 0.944 |
| Knees | 9(28.1%) | 0.900 | 11(26.2%) | 0.531 | 20(33.3%) | 0.513 | 16(34.8%) | 0.476 | 5(38.5%) | 0.725 |
| Ankles/Feet | 10(31.3%) | 0.640 | 9(21.4%) | 0.378 | 17(28.3%) | 0.821 | 15(32.6%) | 0.278 | 3(23.1%) | 1.000 |

NA: Not applicable

## Discussion

In the present study, the overall prevalence of the LMSDs among BMSs was 77.1%. This finding is higher than a similar two studies, one was in Iran, which reported 72.4% MSDs among medical laboratory workers (11). The other one was in India which found that the overall prevalence of MSDs among medical laboratory technicians was 73.3% (8). However, this finding is lower than a recent study in Saudi Arabia which reported 82% MSDs among medical laboratory workers (12).

The most prevalent LMSDs among BMSs were neck followed by shoulders and lower back with 50.6%, 49.4%, 43.4%, respectively. This finding is in line with other study conducted in Metropolitan Washington, which found that neck complaint (61.5%) was the most prevalent symptom. However, their study also found that the wrists/hands (56.4%) symptoms were also high among their participants. This could be because their study only involved cytotechnologists who deal more with microscopes, and frequently use their hands/wrists to adjust the microscope knob (13). Similar finding also reported that neck (55-60%), followed by upper back (53%), and lower back (57%) were the most prevalent MSDs among cytotechnologist (14). In addition, similar findings were reported among laboratory technicians in which they found that neck was the most prevalent MSDs (3,15,16). Another two studies, which used pathologists as a target health professionals found that neck was the most prevalent MSDs (17,18).

In contrast with our findings, a similar study conducted in India, which included medical laboratory technicians from different departments such as pathology, microbiology haematology, histology, and biochemistry. They found that the lower back complaint (32.5%) was the most prevalent followed by knees (20.7%) (19). Another similar study was conducted in Ethiopia revealed that ankles/feet (21.7%) and knees (20.8%) were the most affected parts (3). In addition, two similar studies in Sadia Arabia and Iran found that lower back was the most prevalent MSDs among clinical laboratory workers (11,12).

The finding of our study could be explained by the fact the BMSs used to work with different procedures and deal mostly with specimens that require the use of microscopes. Therefore, they used to bend their neck for a long time. Further, they used to work with static or repetitive tasks without relaxing their arms. This may explain the reasons for their shoulders complaints. The high prevalence of lower back complaints could be due to nonadjustable chairs with unsupported back posture available in their workstations. Another factor that might be a reason for the lower back complaint is the job nature, in which the BMSs used to work for a long time while they were standing without back support. BMSs in the current study reported that their symptoms of LMSDs occurred irregularly (Figure2). This finding is also consistent with other similar study (13).

Some risk factors such as increasing age of medical laboratory technicians, and worsening of the working conditions as well as standing and sitting for long hours were found to be associated with developing MSDs among the medical laboratory workers (3). The current study found no statistically significant association with age, marital status, nationality, dominant hand, BMI, degree of qualifications, and working overtime. Similar findings were also reported (8). Usually the BMSs work in multitasking jobs in which they move from one workstation to another. Therefore, they were able to avoid postural stress. So, this might explain the reasons for no significant association between age and the MSDs (8).

The female gender showed a significant association with neck and upper back symptoms among BMSs (Table 1). This finding is in agreement with other similar study (8). In contrast, other study found that the female gender had no association with a neck complaint (11). In general, the prevalence of MSDs was found to be more common among women rather than men (20). A study conducted in Amsterdam also found that female workers are at high risk to develop MSDs in the upper extremity (21). There were several explanations for this gender difference. One of these explanations is the biological differences between females and males. For example, both genders can expose to the same physical work demands, however, they have different maximal physical work capacities. In addition, many workstations or tools are not taking into account the anthropometric differences between the two genders. Furthermore, it is suggested that the female reproductive system and, use of oral contraceptives, pregnancy, and childbirth can explain the high risk of MSDs among females. However, the precise mechanism is still not explored (21).

In the current study, pipetting showed a significant association with shoulders and lower back complaints. This is consistent with other study that found that increasing pipette work was associated with an increase in shoulder complaints among laboratory technicians (22). The significant association between pipetting and lower back complaints can be explained by the fact that BMSs did not use back supports while pipetting. Furthermore, our finding showed a significant association between microscopy and shoulder complaints. This may due to increased stress of the shoulder ligaments while using the microscope for a long time.

The present study found that there was a significant association between the LMSDs specifically the lower back complaints and the heavy work at home. This was in agreement with similar study in which heavy work of home was also significant with MSDs but with wrists/hands (11). However, both studies found that there was no significant association between stress at work and MSDs among medical laboratories.

Several limitations of our study are worth noting. First, due to the COVID-19 pandemic, we were not able to spread the questionnaire face to face to the BMSs. Therefore, we were incapable to provide more explanation about questions for the participants and encourage them to participate. Second, the study was restricted to BMSs from a single hospital and university, even though this hospital serves as a tertiary referral hospital in Oman and the university is the national university of the Sultanate of Oman. Finally, the study did not distinguish if those LMSDs were partially or fully related to the instruments and machines used in different laboratories.

In conclusion, the study highlighted that the prevalence of LMSDs among BMSs is considerably high. The most affected parts were the neck, followed by the shoulder and lower back. The study also found that the prevalence of LMSD was associated with female gender, pipetting, microscopy, and heavy work at home. Good knowledge, attitude, practice, and training of ergonomics may minimize the prevalence of LMSDs among BMS.

## Data Availability

All data referred to this manuscript are available.

## Acknowledgments

The authors would like to thank all BMSs who participated in this study.

